# Cyproterone Acetate and risk of Meningioma

**DOI:** 10.1101/2020.12.29.20248395

**Authors:** Anders P. Mikkelsen, Iben K. Greiber, Nikolai M. Scheller, Malene Hilden, Øjvind Lidegaard

## Abstract

Cyproterone acetate (CPA) is a synthetic steroid hormone. We assessed the association between the use of CPA and the risk of developing meningioma.

In a historical prospective cohort study, using Danish national healthcare registers we included a cohort of 5,730,654 individuals, among whom 1,982 were exposed to CPA. During follow-up, we identified 8,957 cases of meningioma, of which 16 were exposed to CPA. From 2013 to 2019 the number of new users increased from 18.1 to 62.3 new users per million, while the proportion of new users who were transgender increased from 18.4 to 68.3%. Analyses showed a significantly increased risk of meningioma according to cumulative dose of CPA; 0.1-10 grams of CPA, incidence rate 78.8 (95% CI 15.7-141.9) per 100.000 person years and adjusted hazard ratio 7.0 (3.1-15.6); >10 grams of CPA, incidence 187.5 (71.3-303.7) and adjusted hazard ratio 19.2 (10.3-35.8), as compared to the background population.

In conclusion, the cumulative dose of CPA was associated with an increased incidence and hazard ratio of meningioma, showing a dose-response relationship. The number of new CPA users increased more than 3-fold from 2013 to 2019, primarily driven by new transgender users.

## Introduction

A meningioma is a tumour of the meninges that surrounds the brain and spinal cord. It is the most frequent type of primary brain tumour, with an age-adjusted incidence of 8.56 per 100.000 person years and over 31,000 events in the United States per year.^1^ Meningiomas are slow-growing, and in most cases non-malignant, although they may induce severe morbidity and mortality due to pressure on surrounding structures. Consequently, it is recommended that, if possible, symptomatic meningiomas be surgically removed.

Meningiomas have been associated with advancing age, female sex, neurofibromatosis type 2 and ionizing radiation.^2^ Moreover, the use of the drug cyproterone acetate (CPA) has been associated with the development of meningiomas. CPA may potentially affect meningioma growth due to its anti-androgen and pro-progestin properties, as 2/3 meningiomas express progesterone and androgen receptors.^3,4^ The medication is used by both males and females, although for varying indications. Among individuals of the male sex, a high dosage formulation is used to treat prostate cancer, hypersexuality disorder, and in male-to-female gender affirming hormone therapy. In individuals of the female sex it is used in the treatment of acne, hirsutism, seborrhoea, hair-loss, and in a low-dose variant in combination with ethinylestradiol as birth control.^5,6^

Since 2008, past or present meningioma has been a contraindication to CPA treatment. This was initially based on data from case series, highlighting a potential association between CPA and meningiomas.^7^ Subsequently, two cohort studies found an increased risk of meningioma among users of high-dose CPA, as compared to non-users (odds ratio of 6.3, 95% confidence interval [CI] 1.4-28.9; rate ratio 11.4 95% CI 4.3-30.8), however both studies included less than 5 exposed cases.^8,9^ A recent French cohort study, comparing females exposed to high cumulative doses versus low cumulative doses of CPA, found a 6-20 fold increased risk of meningioma. As a consequence, the European Medicines Agency’s recently issued a recommendation to restrict the use of more than 10 milligrams CPA per day.^10^

In a nationwide register-based cohort study we assessed national trends in use of CPA and the risk of meningioma according to cumulative exposure to CPA, as compared to both non-users among males and females. Furthermore, we assessed if the risk of meningioma among past users of CPA returned to the baseline risk in the general population.

## Materials & methods

### Study design

Historical prospective cohort study.

### Study population

Using the nationwide Danish health and demographic registers we identified all individuals born between 1930 to 2000, using data from the Central Persons Register. Individuals were included in the study on their 15^th^ birthday, on the day of inclusion in the Central Persons Register, or on January 1^st^, 1995, whichever came last. Study participants were then followed until a diagnosis of meningioma, neurofibromatosis type 2, death, emigration, or December 31^st^, 2017, whichever came first. We excluded individuals with prior meningioma, intracranial surgery, neurofibromatosis type 2, or who died before inclusion. Meningioma was the event of interest and defined as a primary or secondary diagnosis in the Danish National Health Register or the Danish Cancer Register. For a summary of diagnosis codes used see Table 1.

**Table 1:** Definitions. Search strings used for diagnosis definitions. Only primary and secondary diagnosis codes from the National Patient Register (LPR) were used, and emergency room contacts were not used. ICD-7: International Classification of Diseases 7^th^ revision. ICD-8: 8^th^ revision. ICD-10: 10^th^ revision. ATC: Anatomical Therapeutic Chemical Classification. [Brackets]: Indicate an interval of codes, both characters included. All subdiagnoses were included. Registers used: CR: Cancer Register, available since 1942. CPR: Central Persons Register, available since 1968. DAR: Cause of Death Register, available since 1969. LPR: National Patient Register, available since 1977. LMDB: National Prescription Register, available since 1995.

| Definition | Registers | ICD-7 | ICD-8 | ICD-10 | Other |
| --- | --- | --- | --- | --- | --- |
| Cyproterone acetate | LMDB |  |  |  | ATC: G03HA01 |
| Oestrogen therapy | LMDB |  |  |  | ATC: G03C |
| Meningioma | LPR, CR | 2930, 2931, 4932 | 22599, 19219, 19239, 2250[0-3], 22507, 22509, 2252[0-9], 22599, 23839 | C70, D32 |  |
| Intracranial surgery | LPR |  |  |  | NCSP: KAA, KAB |
| Prostate hypertropia or cancer | LPR, CR | 1770, 4770, 6770, 7770, 8770 | 6000[0-9] | N40, N510 |  |
| Neurofibromatosis type 2 | LPR |  | 31[0-5]30, 74349 | Q850 |  |
| Hirsutism, acne, or hair-loss | LPR |  | 7040[0-1] | L70, L68, L64, L65, L66 |  |
| Transgender | CPR, LPR |  | 30239, 30259 | F64, Z768E | Gender change in CPR |
| Hypersexuality | LPR |  | 30229, 30289, 30299 | F65 |  |

Information about prescriptions fulfilled for high-dose CPA since January 1^st^, 1995 was extracted from the Danish Prescription Register, using the Anatomical Therapeutic Chemical Classification System (ATC) code G03HA01. CPA was available in the following formats during the study period: (i) tablet of 50 milligrams CPA in packets of 45, 50, 225, or 250 pieces for oral use, (ii) injections with 3 millilitres (100 milligrams/millilitre) CPA for intramuscular use, in packets of 3 pieces. The dose per tablet or injection was multiplied by the number of pieces in each packet, and again multiplied by the number of packets dispensed per prescription. This was used to calculate the dose consumed or injected per prescription, and the cumulative dosage was summed over calendar time. Follow-up was then categorized into three exposure groups (i) No CPA, (ii) 0.1 to 10 grams of CPA, (iii) above 10 grams of CPA.

### New users

We identified new users by the first prescription of CPA from 1997 to 2019 in the Danish Prescription Register with no history of prior use of CPA. The number of new users were then aggregated across each calendar year and divided by all living individuals above 14 years of age in each calendar year. The indication for CPA use was divided into (I) *prostate disease* if new users had a diagnosis of prostate cancer or hypertrophy, or (II) *transgender* if they did not have prostate disease, and fulfilled one or more of the following criteria: (i) changed gender in the Central Persons Register, (ii) relevant diagnosis code, (iii) male sex and fulfilled at least one prescription for oestrogen therapy in addition to CPA, or (iv) male sex, stated indication for the CPA was hormone therapy, and the age below 65 years of age. New users not fulfilling these criteria were grouped as (III) *other*. For the years 2018 to 2019, only data from the National Prescription Register, and about transgender changes in the Central Persons Register were available.

### Statistical analyses

Using a time-dependent Cox regression model, hazard ratios were estimated using age as the underlying time scale. Risk time started at age of inclusion, covariates were updated at the ages were changes occurred, and ended at the age of censoring. To account for confounders, estimates were adjusted for calendar year in 5-year intervals (spline with 4 knots), maternal age (numerical), history of parental meningioma, and sex. The proportional hazard assumption was tested by plotting the Schoenfeld residuals over time with no sign of violation.

### Past or present use

In a separate analysis, we characterized users according to non, present, or past use of CPA. In this analysis, the study population was initially considered as non-users of CPA. At the date of the first prescription for CPA, individuals were considered current CPA users, and 365 days after the last prescription for CPA, individuals were considered past users.

Software used for data management and statistical analyses was *R* version 4.0.2^11^, and packages *data*.*table* version 1.13.0^12^, and *survival* version 3.2-3^13^.

## Results

A total of 5,730,654 individuals were eligible for inclusion in the cohort study and 1,982 (0.03%) were treated with CPA at some time during follow-up. As individuals could contribute with person years in each cumulative exposure group, 1,977 individuals were exposed to 0.1 to 10 gram of CPA, and 781 individuals were exposed to >10 gram of CPA, see Fig. 1. Among users of CPA 1,729 (87.2%) were males, 1,055 (53.2%) had prostate disease, and 204 (10.3%) had a transgender diagnosis or had their gender changed in the Central Persons Register. The mean follow-up time was 16.5 years. The number of new users of CPA in Denmark increased from 18.1 new users per million people per year in 2013 to 60.7 in 2019, see Fig. 2. The proportion of new users of CPA classified to be transgender increased from 18.4% in 2013 to 68.3% in 2019.

**Figure 1.**
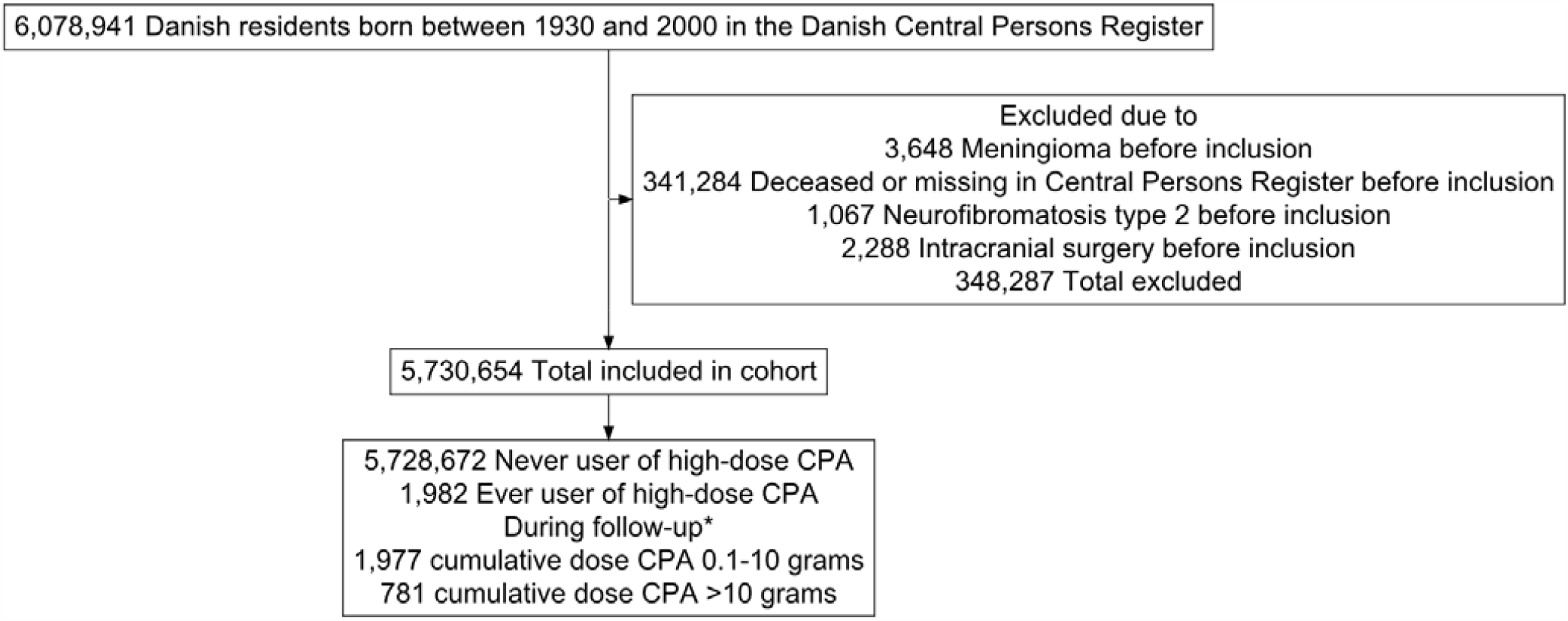
Study inclusion and exclusion criteria. *As exposure to cyproterone acetate (CPA) was time-dependant, the n reflects the largest number of exposed during follow-up.

**Figure 2.**
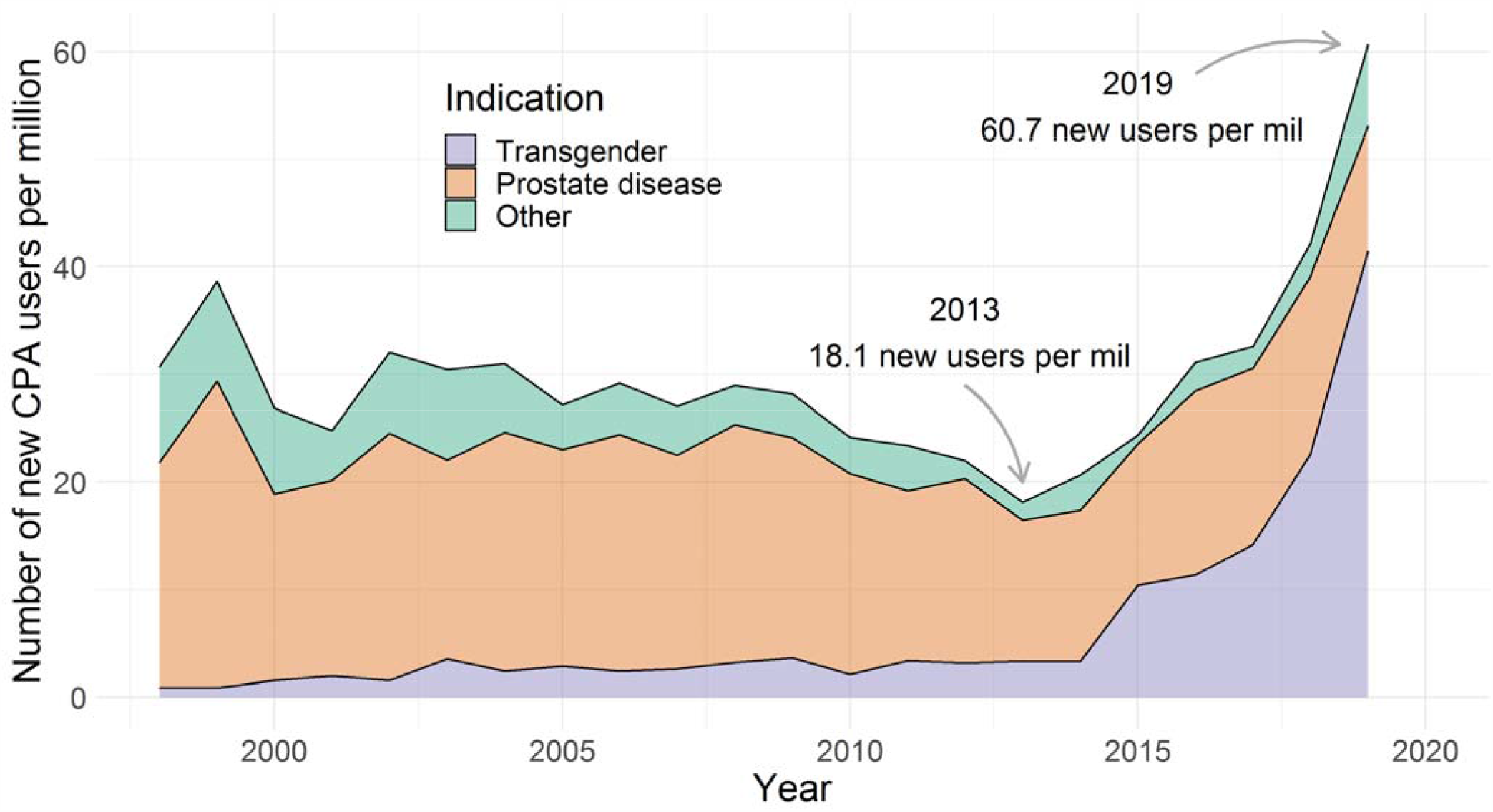
The number of new users of cyproterone acetate (CPA) in Denmark from 1997 to 2019 per million persons alive and above 14 years of age. New users were stratified by indication. mil: million persons.

Individuals not exposed to CPA experienced 8,941 meningioma events in 95,983,527 person years, corresponding to an incidence of 9.3 per 100.000 person years (95% CI 9.1-9.5). Those exposed to 0.1 to 10 grams of CPA experienced 6 meningiomas in 7,610 person years, incidence 78.8 per 100,000 person-years (15.7-141.9), and adjusted hazard ratio was 7.0 (3.1-15.6), as compared to non-users. Persons exposed to over 10 grams of CPA experienced 10 events in 5,333 person years, corresponding to an incidence of 187.5 (71.3-303.7) per 100,000 person years, and an adjusted hazard ratio of 19.2 (10.3-35.8), as compared to non-users, see Table 2a.

**Table 2:** Incidence and hazard ratios of meningioma by use of Cyproterone acetate. Upper part: event rates and hazard ratios (HR) of meningioma by cumulative dose of Cyproterone acetate (CPA). Lower part: Event rates and hazard ratios of past or present use of CPA, compared to non-users. Users were considered past users 1 year after the last prescription of Cyproterone acetate. *Estimates adjusted for age, calendar year in 5-year intervals, parental history of meningioma, and sex. CI: Confidence interval. g: grams.

| Cumulative CPA dose | Events | Person years | Event rate per 100.000 person years (95% CI) | Crude HR (95% CI) | Adjusted HR (95% CI)* | Plot adjusted HR |
| --- | --- | --- | --- | --- | --- | --- |
| None | 8,941 | 95,983,527 | 9.3 (9.1–9.5) | 1 | 1 | • |
| 0.1–10g | 6 | 7,610 | 78.8 (15.7–141.9) | 5.2 (2.4–11.6) | 7.0 (3.1–15.6) | —•— |
| >10g | 10 | 5,333 | 187.5 (71.3–303.7) | 13.1 (7–24.3) | 19.2 (10.3–35.8) | —•— |

*Table 2. Upper part: event rates and hazard ratios (HR) of meningioma by cumulative dose of Cyproterone acetate (CPA).
| CPA use | Events | Person years | Event rate per 100.000 person years (95% CI) | Crude HR (95% CI) | Adjusted HR (95% CI)* | Plot adjusted HR |
| --- | --- | --- | --- | --- | --- | --- |
| Non-users | 8,941 | 95,983,527 | 9.3 ( 9.1– 9.5) | 1 | 1 | • |
| Past | 6 | 8,063 | 74.4 (14.9–133.9) | 5.1 (2.3–11.4) | 5.4 (2.4–12.3) | —•— |
| Present | 10 | 4,881 | 204.9 (77.9–331.9) | 13.6 (7.3–25.2) | 16.0 (8.5–30.4) | —•— |

Past users of CPA, regardless of cumulative dose, had an adjusted hazard ratio of meningioma of 5.4 (2.4-12.3), compared to non-users, see Table 2b.

## Discussion

In a large nationwide Danish register-based study with up to 22 years of follow-up, we found that cumulative dose of CPA was significantly associated with the development of meningioma. A cumulative dose of 0.1 to 10 grams of CPA was associated with a 7-fold increased risk, and cumulative doses above 10 grams was associated with a 19-fold increased risk of meningioma, as compared to non-users. Past users of CPA were still associated with a 5-fold increased risk of meningioma one year after their last prescription for CPA, indicating that the risk declines after cessation but may not return to baseline. Additionally, the incidence of new users increased by more than three-fold, driven largely by new transgender male-to-female users.

### Strengths and weaknesses of the study

The strengths of the study include the nationwide design, prospectively collected data, registered on an individual level, which minimizes the risk of selection bias. Although the cohort was large, treatment with CPA was very rare in the general population, with only 0.03% fulfilling a prescription during the study period. We suspect that a growing tendency to buy medicines online without a doctor’s consent, and at prices often lower than those in pharmacies, the actual number of CPA users may be much higher. This could be diluting the effect of users compared to non-users. However, it can be speculated that this effect may be especially pronounced in 0.1 to 10-gram cumulative CPA group, and that some individuals may have taken a higher dose than registered, overestimating the effect of a certain dose. A 2014 study from the United Kingdom showed that one in four newly referred birth-assigned males self-prescribed hormones before attending a gender identity clinic, most often sourced online.^14^

A French study from 2020 by Weill A. et al.^15^ examined a female population of over a quarter million individuals who were prescribed CPA. Of these women, 516 were subsequently diagnosed with meningioma. They compared groups of women prescribed increasing cumulative doses of CPA, with women prescribed less than three grams of CPA cumulatively. They found an incidence of meningioma after exposure to over 60 grams of CPA of 206 per 100.000 person-years, and an adjusted hazard ratio 21.7 (95%CI 10.8-43.5). Despite differences in the study design, their findings are in accordance with ours. However, our study further adds that even after a low cumulative dose of CPA (i.e., under 10 grams, corresponding to <200 tablets of 50 milligrams) individuals are at 7-fold increased risk of meningioma, and that the risk extends beyond female users, males are equally affected.

An interesting aspect about meningiomas diagnosed after CPA exposure is that they may differ from other meningiomas. They are more often located in the anterior part of the skull base.^15^ Further, CPA meningiomas appear to stagnate or regress after cessation of CPA exposure.^16^ This should warrant neurosurgeons to assess past or present use of CPA when treating patients with meningioma, discontinue the medication, and possibly promote watchful waiting as a first line measure.

## Conclusion

In conclusion, our study found CPA use is strongly associated with later development of meningioma, and the use of CPA is currently rising rapidly mainly due to new transgender users. The association between CPA and meningioma has now shown reproducibility in two large cohort studies, with a clear dose-response effect, large measures of association, and statistical significance. Further, the physiological mechanism is seemingly plausible, with CPA effecting androgen and progestin receptors of tumour precursors, although this was not investigated in the current study. As a randomized controlled trial examining this effect would likely be deemed unethical, available evidence points to a causal relationship between the use of CPA and the development of a meningioma. These results should warrant policymakers to further restrict the use of high dose CPA for non-life threating diseases, as alterative antiandrogens and progestins exist without such potentially lethal side effects. Future research may well examine the effects of other exogenous hormones on the risk of meningioma.

## Data Availability

Data was made available from the Danish Health Data Authority under license FSEID-00003879 and hosted by Statistics Denmark.

## Funding

The current investigation was funded by a grant from The Research Fund of Rigshospitalet, Copenhagen University Hospital grant number E-22515-01. The funders had no role in the design of the study, in the collection, analysis, and interpretation of the data, in the writing of the report; or in any decision related to publication.

## Competing interests

The authors report no competing interests.

## Abbreviations

ATC: Anatomical Therapeutic Chemical Classification System
CI: Confidence interval
CPA: Cyproterone acetate
HR: Hazard ratio

## Notes

### Competing Interest Statement

The authors have declared no competing interest.

### Author Declarations

Approval was granted by the Danish Health Data Authority under license FSEID-00003879 and the Danish Regional Data Protection Agency license P-2020-217.

